# Medical care needs and experiences of LGBTQ populations in Japan

**DOI:** 10.1101/2024.05.30.24308201

**Authors:** Hiroyuki Otani, Tatsuya Morita, Hongja Kim, Kaori Aso, Misuzu Yuasa, Hideyuki Kashiwagi, Kiyofumi Oya, Akemi Shirado Naito

## Abstract

This study comprehensively examines the medical needs and experiences of the lesbian, gay, bisexual, transgender/transsexual, and queer/questioning (LGBTQ) population—also referred to as sexual minorities—in Japan. It aims to bridge the existing gap in understanding the experiences of LGBTQ populations in accessing healthcare, and inform future healthcare reforms. In November 2022, a cross-sectional, web-based, anonymous survey was conducted targeting LGBTQ populations across Japan who had previously visited a medical institution. Participants were recruited through a private, web-based, survey company. Inclusion criteria included being 20 years old or above, having a record of medical visits, and experiencing distress or discomfort related to gender identity, gender, or sexual orientation. Survey items were developed based on previous research and preliminary interviews, to assess demographic characteristics, experiences with medical care, and preferences for end-of-life care. A total of 103 respondents with a diverse demographic profile from across Japan participated in the survey. Among sexual minorities whose gender identity differed from their birth assignment, significant challenges were reported, including distressful experiences related to assigned hospital rooms and difficulties accessing certain medical departments. LGBTQ individuals with non-heterosexual orientations also faced barriers to partner involvement in medical decision-making and care. This study underscores the need for healthcare reforms to address the challenges faced by LGBTQ individuals in Japan. Healthcare providers should create a more equitable and affirming healthcare system for all individuals, irrespective of sexual orientation or gender identity.

## Introduction

The lesbian, gay, bisexual, transgender/transsexual, and queer/questioning (LGBTQ) community is also referred to as sexual minorities [1]. Previously published surveys estimate that approximately 3%–12% of adults in the U.S. population identify as LGBTQ [2].

Previous studies have highlighted the pervasive disparities faced by LGBTQ patients, including higher incidence of certain health conditions, such as mental health disorders, most commonly anxiety and depression [3], compared with their heterosexual and cisgender counterparts [2]. Despite these alarming trends, little attention has been paid to LGBTQ individuals in the field of medical healthcare [4, 5], leading to many cases of patients becoming seriously ill owing to hesitance in seeking medical attention, fearing that medical personnel may not understand their needs [2]. Historically, LGBTQ individuals have encountered numerous barriers to accessing quality healthcare stemming from societal stigma, discrimination, and a lack of understanding within the medical community [2]. Patients have perceived certain healthcare professionals as openly homophobic or harboring unconscious biases regarding sexuality and gender that are either incorrect or offensive [6]. Additionally, 1 out of every 4 LGBT individuals encounters inappropriate curiosity from healthcare professionals owing to a lack of comprehension, while 1 in 8 experiences differential treatment from healthcare providers because of their LGBT status. Furthermore, 1 in 7 individuals within the LGBT community refrains from seeking treatment because of concerns regarding facing discrimination [7]. This background has led to the publication of various guidelines and best practices for the consideration of LGBTQ patients [2, 8-10]. However, despite these efforts, LGBTQ populations continue to face considerable challenges in accessing culturally competent and inclusive care, particularly in regions where societal attitudes toward them remain entrenched in prejudice and discrimination.

In Japan, societal attitudes toward LGBTQ individuals have been a subject of international scrutiny and criticism. While progress has been made in recent years, particularly with the passage of anti-discrimination laws and recognition of same-sex partnerships in certain municipalities, currently, same-sex marriage is not legally recognized in Japan. There remains a pervasive cultural reluctance to openly address issues related to sexual orientation and gender identity [11].

LGBTQ populations have also experienced marginalization within the medical field. For instance, the lack of scholarly attention to the healthcare needs of LGBTQ older adults in Japan [12] highlights a critical gap in our understanding of the unique challenges faced by this demographic. Similarly, the dearth of LGBTQ-inclusive education within Japanese medical schools [13, 14] underscores the need for systemic reforms to ensure that future healthcare professionals are equipped to provide competent and affirming care to LGBTQ patients.

This situation has resulted in a lack of empirical data regarding the experiences and needs of LGBTQ patients in Japan [12]. This has created a significant gap in our understanding of the experiences of LGBTQ patients. This study aims to address this gap by comprehensively examining the medical and care needs and experiences of LGBTQ patients.

## Methods

### Study design and setting

In November 22, 2022, we conducted a cross‐sectional, web-based, anonymous nationwide survey of LGBTQ populations in Japan with a history of visiting a medical institution.

### Participants and procedure

Participants were recruited through a private, web-based, survey company (MACROMILL; Tokyo, Japan). The inclusion criteria were as follows: (a) being 20 years of age or older; (b) having a history of visiting a medical institution; (c) To identify sexual minority (LGBTQ) individuals, respondents were asked the question based on previous research [3, 15], “Have you ever experienced distress or discomfort or dysphoria regarding your physical gender, gender identity, or sexual orientation?” Those who answered “yes” were identified as “sexual minority (LGBTQ).” The survey company recruited potential participants across Japan through convenient sampling and sent questionnaires to them online. Responses to the questionnaire were deemed as consent to participate. Participation was voluntary and confidentiality was maintained throughout all investigations and analyses. The participants received a small reward from the survey company for completing the questionnaire, and no follow‐up was required after the survey completion. We chose MACROMILL as the market research company based on previous research [16, 17].

### Measurements

Survey items were developed based on previous research [9, 10] and preliminary interviews to explore needs and experiences with medical care, including issues in the hospital environment and key personnel related to the LGBTQ populations.

#### Demographic and clinical characteristics

Data on demographic and clinical characteristics were obtained from the self-reported questionnaires. The data included: 1) age; 2) gender ([i] gender assigned at birth, [ii] gender identity: What gender do you identify yourself as?, and [iii] sexual orientation: Which genders are you sexually attracted to?); and 3) marital status.

#### Questions regarding medical care for LGBTQ populations

We surveyed individuals identifying as LGBTQ about their experiences with medical care using a 5-point scale ranging from “not distressful” to “very distressful” along with the free-text section. Specifically, respondents shared instances such as, “I found it challenging to visit the outpatient clinic owing to discomfort with being addressed by my name,” “It took me a long time to see a doctor because I was worried that I would have a bad experience regarding sexual matters,” “It was difficult to visit departments with a strong sexual impression, such as gynecology and urology,” “The doctor or nurse approached me based on my external gender (e.g., do you have a boyfriend/girlfriend),” “I encountered negative comments about my gender identity or sexual orientation from doctors or nurses,” “In the hospital, I was assigned to a room of a gender different from the one I identify as,” “I had to use a toilet that was labeled for a different gender from the one I identify as,” “It was difficult to ask a nurse or caregiver for assistance with toileting,” “It was difficult to ask nurses or caregivers for assistance with changing clothes or personal-hygiene tasks (showering, wiping, etc.),” and “I wanted to talk to other patients who were experiencing similar challenges but couldn’t bring myself to do so.”

Additionally, individuals with partners of a non-heterosexual sexual orientation were asked to rate the following experiences on a 5-point scale ranging from “not distressful” to “very distressful”: “I wanted to discuss my medical condition with my partner but could not,” “I wanted my partner to be involved in deciding my treatment plan with me but could not,” “I could not introduce my partner as my partner (had to introduce my partner as a friend),” “I was required to have an explanatory consent form signed by a family member and was informed that my partner’s consent was not acceptable (the hospital required consent from a blood relative),” “In deciding on the treatment plan, the opinions of blood relatives were assigned more weight than those of my partner,” “During hospitalization, I was unable to obtain permission for my partner to visit,” and “My partner could not accompany me during surgery.”

#### Questions regarding participants’ wishes for medical institutions during end of life

Participants were surveyed using a 5-point scale, ranging from “not important” to “essential,” regarding their preferences for medical institutions during end of life. Specifically, respondents were asked to rate the importance of the following factors: “Being able to be hospitalized in a room corresponding to one’s gender identity,” “Opening up to medical personnel about one’s gender identity and receiving appropriate support,” “If you have a partner, you can have your medical condition explained to both you and the partner, with the partner receiving the information on your behalf as a family member,” “If you have a partner, you are allowed to meet with them and stay together overnight,” and “If you have a partner, they can be present at your deathbed.”

### Statistical analyses

Broadly, sexual minorities (LGBTQ) are defined as individuals who have experienced concerns or discomfort regarding their physical, mental, or sexual orientation. The following question was used in a study conducted in March 2015 to identify sexual minority individuals: “Have you ever experienced distress or discomfort or dysphoria regarding your physical gender, gender identity, or sexual orientation?” [3, 15]. Additionally, a statistical analysis was performed defining those whose gender identity differed from their assigned gender at birth and those whose sexual orientation diverged from heterosexuality [2, 3, 15]. Descriptive statistical analysis was employed as the analysis methodology. This involved computing the frequency of responses such as “a little distressful,” “distressful,” and “very distressful” on a 5-point scale ranging from “not distressful” to “very distressful.” Similarly, the frequency of responses indicating importance, ranging from “not important” to “essential,” including “important” and “very important,” was also computed. This is an exploratory descriptive study; the required number of cases for the expected frequency of 20% to have a confidence interval width of 15% was 109. Therefore, the target number of cases for the study was set at 100.

## Results

A total of 103 patients from all 8 regions of Japan responded to the survey. The respondents whose assigned gender at birth, gender identity (currently perceived gender), and sexual orientation (the gender to which they were attracted) were male amounted to 56 (54.4%), 51 (49.5%), and 45 (43.7%), respectively. The most frequent age group (years) was 40–49 (30.0%), with 31 respondents, followed by 50–59 (23.3%), with 24 respondents. Further, 62 (60.2%) of the respondents were married (Table 1). The most frequent response among the 103 people who identified as sexual minorities in a broad sense (LGBTQ) was, “It was difficult to visit departments with a strong sexual impression, such as gynecology and urology” (51.5%; Table 2).

**Table 1.**
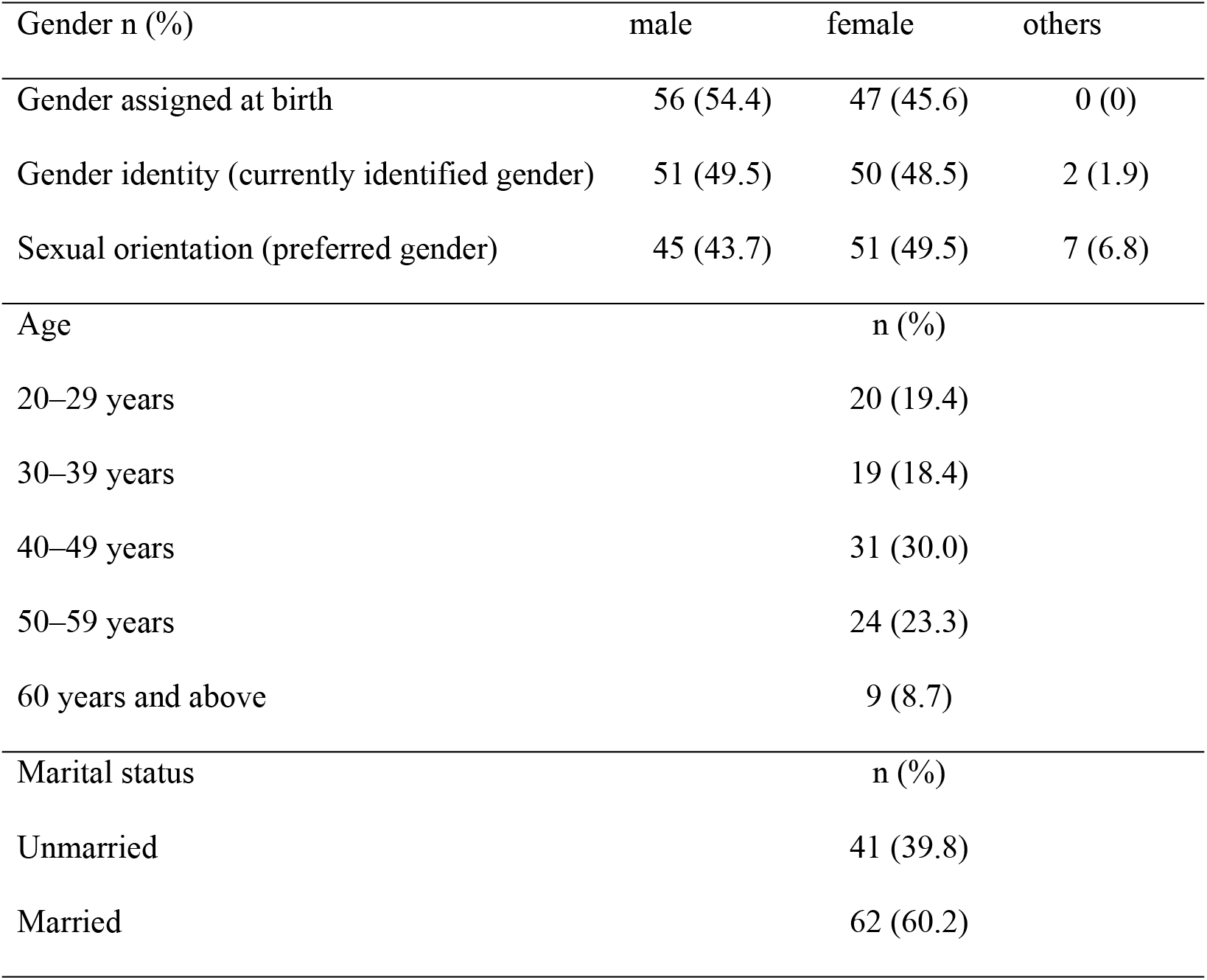
Participants’ characteristics (n=103)

**Table 2.**
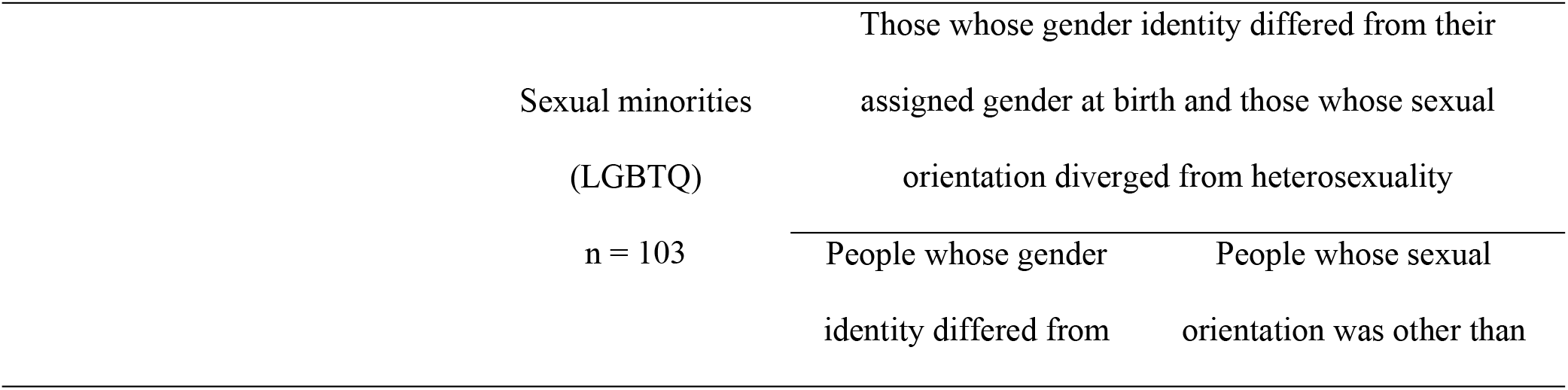
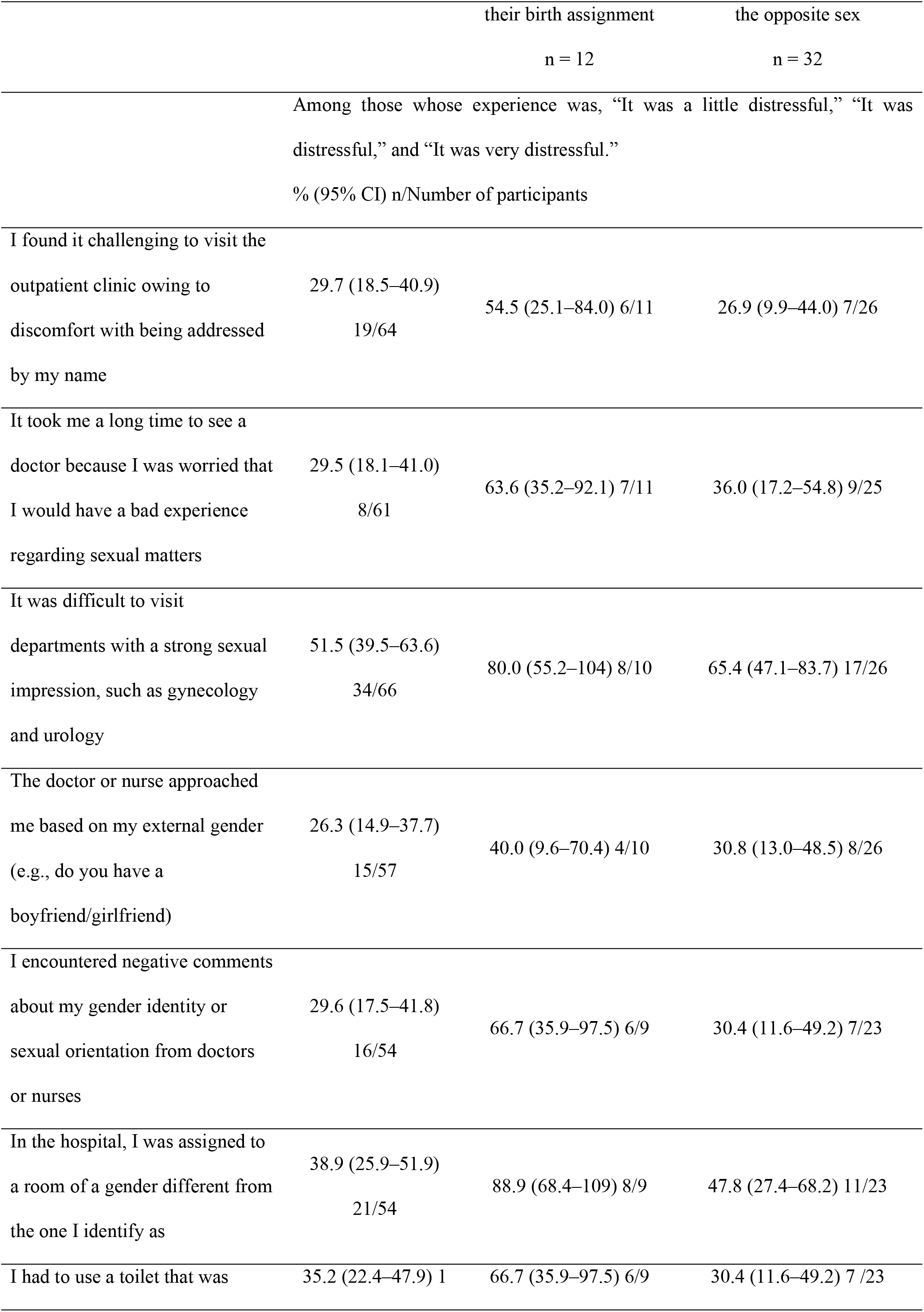
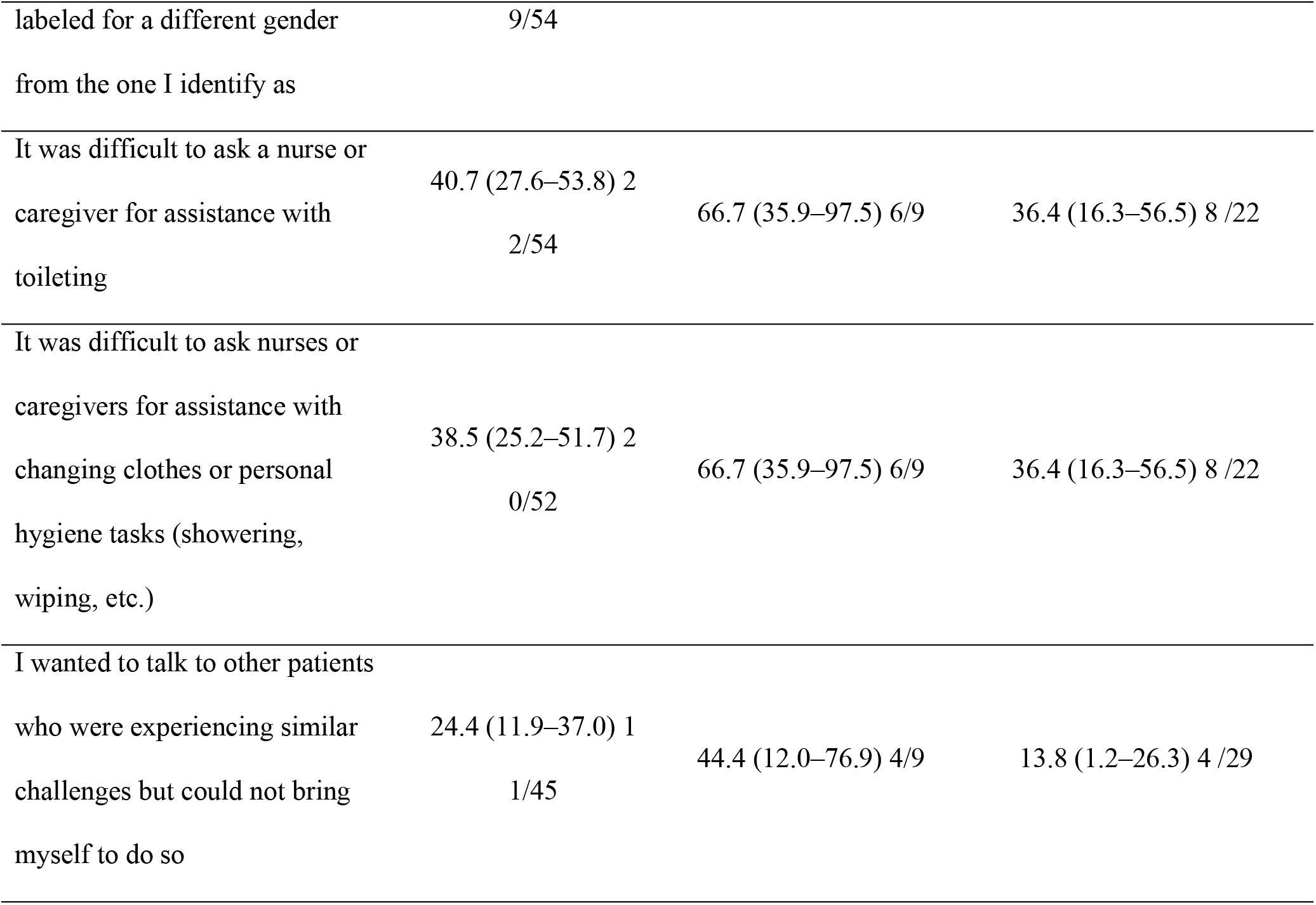
Experience when visiting a medical institution.

### Those whose gender identity differed from their assigned gender at birth and those whose sexual orientation diverged from heterosexuality

Twelve people whose gender identity differed from their assigned gender at birth and 32 people whose sexual orientation was other than the opposite sex were analyzed as sexual minorities (LGBTQ)

#### Those whose gender identity differed from their assigned gender at birth

Among those whose gender identity differed from their assigned gender at birth, the most frequent responses were “In the hospital, I was assigned to a room of a gender different from the one I identify as” (88.9%), and “It was difficult to visit departments with a strong sexual impression, such as gynecology and urology” (80.0%), indicating that many respondents felt that it was distressful to see a doctor. In addition, in the free-text section, participants expressed their hardships, such as, “When my name was called out loud, I did not like the reactions of the people around me, and it thus bothered me” (Table 2).

#### Those whose sexual orientation diverged from heterosexuality

Among those whose sexual orientation diverged from heterosexuality, “My partner could not accompany me during surgery” (58.3%), “I wanted to discuss my medical condition with my partner but could not” (57.1%), and other issues were raised (Table 3).

**Table 3.**
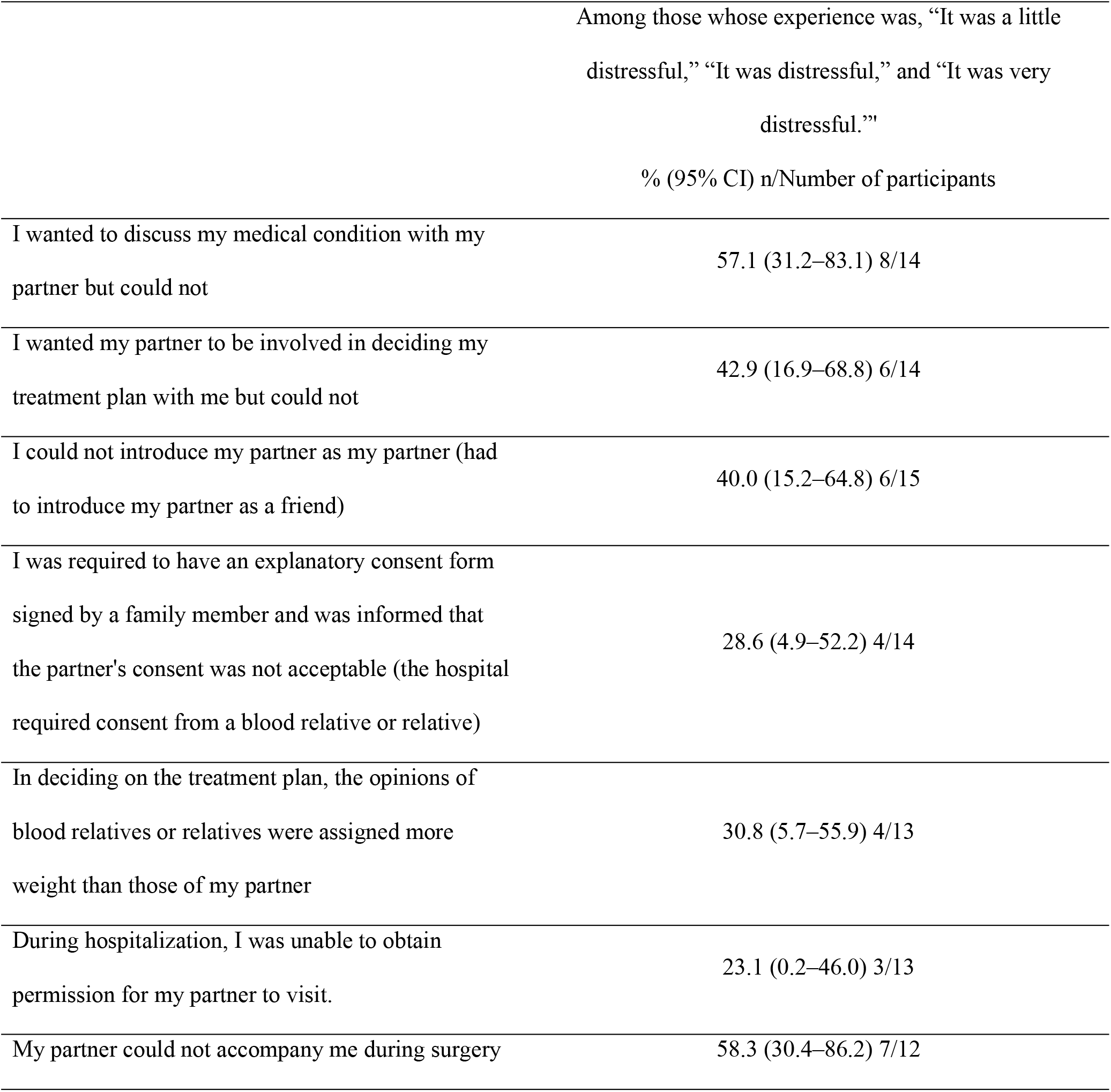
Experiences of individuals with partners of a non-heterosexual sexual orientation at medical institutions (n = 23)

#### Wishes in case of end-of-life stage

Of the respondents, 75.0% and 65.6% of those whose gender identity differed from their assigned gender at birth and those whose sexual orientation diverged from heterosexuality, respectively, indicated “important” in their response to the statement, “If you reach the end-of-life stage, your hope for medical institutions is that if you have a partner, they can be present at your deathbed.” (Table 4).

**Table 4.**
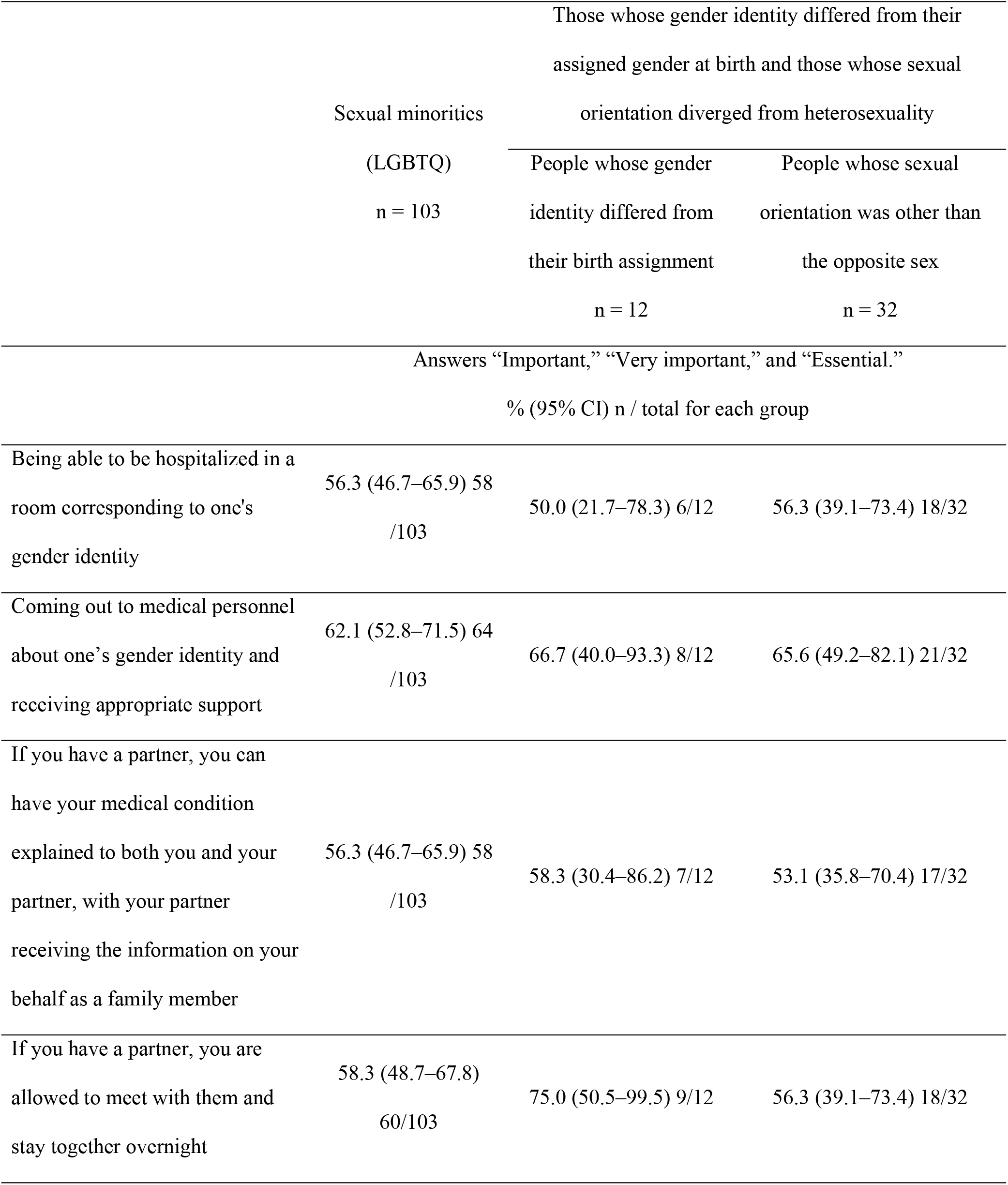
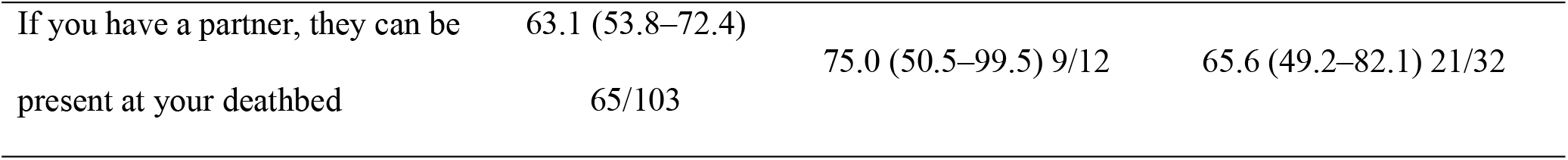
Wishes for medical institutions in case of end-of-life stage.

## Discussion

This cross-sectional, web-based, anonymous study revealed the medical care needs and experiences of the LGBTQ populations in Japan. The findings of this study shed light on the significant challenges faced by LGBTQ individuals in Japan when accessing medical care, particularly regarding their gender identity, sexual orientation, and the inclusivity of healthcare environments. The results underscore the need for healthcare reforms that prioritize cultural competency and sensitivity to the needs of sexual-minority populations.

The experiences reported by sexual minorities reveal systemic challenges within the healthcare system. For instance, the finding that a proportion of respondents felt distress when assigned to hospital rooms based on their assigned gender at birth highlights the need for gender-affirming practices within medical institutions. Similarly, difficulties in accessing certain departments, such as gynecology and urology, owing to their strong sexual impression, indicate the presence of institutional barriers that hinder LGBTQ individuals from seeking necessary medical care. These facts have also been reported in previous studies overseas as adults in the U.S. population who identify as LGBTQ [2], where best practices for the consideration of LGBTQ patients are being considered [2, 8-10].

Moreover, the experiences reported by LGBTQ individuals with non-heterosexual sexual orientations underscore the importance of inclusive policies regarding partner involvement in medical decision-making and care. The inability of partners to accompany respondents during surgery and the challenges in discussing medical conditions with partners highlight the lack of recognition and support for LGBTQ relationships within healthcare settings. Similar experiences have been found in the context of COVID-19, and education to strive toward inclusive person-centered care in sensitive and respectful ways, including legal aspects, is necessary for medical professionals [10].

Furthermore, the preferences expressed by respondents regarding end-of-life care emphasize the significance of inclusive practices that honor individuals’ chosen identities and relationships. The desire for partners to be present at each other’s deathbeds reflects the importance of acknowledging and respecting LGBTQ relationships in the context of medical care, particularly during sensitive and vulnerable stages of life. These wishes are desired by LGBTQ individuals and also among family members of cancer patients [18]. Medical professionals must be consistently considerate of all patients.

This study’s findings contribute to the growing body of research on LGBTQ-healthcare disparities in Japan and underscore the urgent need for systemic reforms. Healthcare providers and policymakers must prioritize LGBTQ-inclusive education and training to ensure that all individuals receive equitable and affirming care, regardless of their sexual orientation or gender identity. Additionally, healthcare institutions should implement policies and practices that promote gender-affirming care, support LGBTQ relationships, and create inclusive environments that foster trust and comfort among LGBTQ patients.

### Strengths and limitations

This study’s strengths include the demographic profile of the respondents that reflects a diverse range of ages and marital statuses, indicating that LGBTQ individuals seeking medical care in Japan represent a broad spectrum of the population. However, this study has some limitations. First, as we applied convenient sampling via the Internet using a private, web-based company and analyzed the first 103 responders, we could not extract a response rate or the characteristics of non-responders. This sampling method may introduce selection bias. Second, we used questionnaires that had not been validated. Future research must address the individuality of each LGBTQ population, including large-scale studies.

## Conclusions

This study highlights the pervasive challenges faced by LGBTQ individuals in accessing quality healthcare in Japan, and underscores the importance of addressing these disparities through comprehensive reforms. By prioritizing cultural competency, inclusivity, and sensitivity to the needs of sexual-minority populations, healthcare providers and policymakers can work toward creating a healthcare system that is truly equitable and affirming for all individuals, regardless of their sexual orientation or gender identity.

## Data Availability

Data cannot be shared publicly because of privacy. Data are available from the St. Mary’s Hospital Institutional Data Access / Ethics Committee (contact via the local Institutional Review Board of St. Mary’s Hospital) for researchers who meet the criteria for access to confidential data.

## Author contributions

Study concept and design: Hiroyuki Otani, Tatsuya Morita, Hongja Kim, Kaori Aso, Misuzu Yuasa, Hideyuki Kashiwagi, Kiyofumi Oya, Akemi Shirado Naito.

Collection and/or assembly of data: Hiroyuki Otani, Tatsuya Morita, Hongja Kim, Kaori Aso, Misuzu Yuasa, Hideyuki Kashiwagi, Kiyofumi Oya, Akemi Shirado Naito.

Statistical analysis: Hiroyuki Otani, Tatsuya Morita, Hongja Kim, Kaori Aso, Misuzu Yuasa, Hideyuki Kashiwagi, Kiyofumi Oya, Akemi Shirado Naito.

Data analysis and interpretation: Hiroyuki Otani, Tatsuya Morita, Hongja Kim, Kaori Aso, Misuzu Yuasa, Hideyuki Kashiwagi, Kiyofumi Oya, Akemi Shirado Naito.

Drafting of the manuscript: Hiroyuki Otani, Tatsuya Morita, Hongja Kim, Kaori Aso, Misuzu Yuasa, Hideyuki Kashiwagi, Kiyofumi Oya, Akemi Shirado Naito.

Final approval of the manuscript: Hiroyuki Otani, Tatsuya Morita, Hongja Kim, Kaori Aso, Misuzu Yuasa, Hideyuki Kashiwagi, Kiyofumi Oya, Akemi Shirado Naito.

## Declarations

### Ethical considerations

This study was conducted in accordance with the ethical standards of the Helsinki Declaration and ethical guidelines for medical and health research involving human subjects presented by the Ministry of Health, Labour, and Welfare in Japan. It was also approved by the local Institutional Review Board of St. Mary’s Hospital (Approval number 22-0804).

### Consent to participate

Responses were considered consent to participate. Responses to the questionnaire were voluntary, and confidentiality was maintained throughout all investigations and analyses.

### Consent for publication

All authors agree to the manuscript submission.

### Data availability

The datasets supporting the study results are available from the corresponding author on reasonable request.

## References

1. Rosser BRS, Weideman BCD, Rider GN, Jatoi A, Ecklund AM, Wheldon CW, et al. Sexual and gender minority invisibility in cancer studies: A call for effective recruitment methods to address cancer disparities. J Clin Oncol. 2023;41(33):5093–5098. 10.1200/JCO.23.00655

2. Quinn GP, Sanchez JA, Sutton SK, Vadaparampil ST, Nguyen GT, Green BL, et al. Cancer and lesbian, gay, bisexual, transgender/transsexual, and queer/questioning (LGBTQ) populations. CA Cancer J Clin. 2015;65(5):384–400. 10.3322/caac.21288

3. Dhejne C, Vlerken RV, Heylen G, Arcelus J. Mental health and gender dysphoria: A review of the literature. Int Rev Psychiatry. 2016;28(1):44–57. 10.3109/09540261.2015.1115753

4. Schabath MB, Blackburn CA, Sutter ME, Kanetsky PA, Vadaparampil ST, Simmons VN, et al. National survey of oncologists at National Cancer Institute-designated comprehensive cancer centers: Attitudes, knowledge, and practice behaviors about LGBTQ patients with cancer. J Clin Oncol. 2019;37(7):547–558. 10.1200/JCO.18.00551

5. Wheldon CW, Schabath MB, Hudson J, Bowman Curci M, Kanetsky PA, et al. Culturally competent care for sexual and gender minority patients at national cancer institute-designated comprehensive cancer centers. LGBT Health. 2018;5(3):203–211. 10.1089/lgbt.2017.0217

6. Malta M. LGBTQ+ health: Tackling potential health-care professionals’ bias. Nat Rev Dis Primers. 2023;9:1. 10.1038/s41572-022-00413-2

7. Webster R, Drury-Smith H. How can we meet the support needs of LGBT cancer patients in oncology? A systematic review. Radiography. 2021;27(2):633–644. 10.1016/j.radi.2020.07.009

8. Fasullo K, McIntosh E, Buchholz SW, Ruppar T, Ailey S. LGBTQ older adults in long-term care settings: An integrative review to inform best practices. Clin Gerontol. 2022;45(5):1087–1102. 10.1080/07317115.2021.1947428

9. Maingi S, Bagabag AE, O’Mahony S. Current best practices for sexual and gender minorities in hospice and palliative care settings. J Pain Symptom Manage. 2018;55(5):1420–1427. 10.1016/j.jpainsymman.2017.12.479

10. Rosa WE, Shook A, Acquaviva KD. LGBTQ+ inclusive palliative care in the context of COVID-19: Pragmatic recommendations for clinicians. J Pain Symptom Manage. 2020;60(2):e44–e47. 10.1016/j.jpainsymman.2020.04.155

11. The Lancet. Coercive sterilisation of transgender people in Japan. Lancet. 2019;393(10178):1262. 10.1016/S0140-6736(19)30739-1

12. Bratt AS, Hjelm A-CP, Wurm M, Huntley R, Hirakawa Y, Muraya T. A systematic review of qualitative research literature and a thematic synthesis of older LGBTQ people’s experiences of quality of life, minority joy, resilience, minority stress, discrimination, and stigmatization in Japan and Sweden. Int J Environ Res Public Health. 2023;20:6281. 10.3390/ijerph20136281

13. Yamazaki Y, Aoki A, Otaki J. Prevalence and curriculum of sexual and gender minority education in Japanese medical school and future direction. Med Educ Online. 2020;25(1):1710895. 10.1080/10872981.2019.1710895

14. Yoshida E, Matsushima M, Okazaki F. Cross-sectional survey of education on LGBT content in medical schools in Japan. BMJ Open. 2022;12(5):e057573. 10.1136/bmjopen-2021-057573

15. Cooper K, Russell A, Mandy W, Butler C. The phenomenology of gender dysphoria in adults: A systematic review and meta-synthesis. Clin Psychol Rev. 2020;80:101875. 10.1016/j.cpr.2020.101875

16. Mori M, Fujimori M, Ishiki H, Nishi T, Hamano J, Otani H, et al. Adding a wider range and “hope for the best, and prepare for the worst” statement: Preferences of patients with cancer for prognostic communication. Oncologist. 2019;24(9):e943–e952. 10.1634/theoncologist.2018-0643

17. Morita T, Kiuchi D, Ikenaga M, Abo H, Maeda S, Aoyama M, et al. Difference in opinions about continuous deep sedation among cancer patients, bereaved families, and physicians. J Pain Symptom Manage. 2019;57(3):e5–e9. 10.1016/j.jpainsymman.2018.11.025

18. Otani H, Yoshida S, Morita T, Aoyama M, Kizawa Y, Shima Y, et al. Meaningful communication before death, but not present at the time of death itself, is associated with better outcomes on measures of depression and complicated grief among bereaved family members of cancer patients. J Pain Symptom Manage. 2017;54(3):273–279. 10.1016/j.jpainsymman.2017.07.010

